# “Holy cow, where do I sign up?” Attitudes of Military Veterans toward Epigenomic Biomarker Toxic Exposure Testing

**DOI:** 10.1101/2024.04.09.24305554

**Authors:** Stacey Pereira, Calvin Apodaca, Kyle Slominski, Rachele K. Lipsky, Cristian Coarfa, Cheryl L. Walker, Amy L. McGuire, Lea Steele, Drew A. Helmer

## Abstract

**Background:** With the signing of the PACT Act in 2022, there is great interest and investment in studying toxic exposures encountered during military service. One way to address this is through the identification of epigenomic biomarkers associated with exposures. There is increasing evidence suggesting that exposure to toxic substances may result in alterations to DNA methylation and resultant gene expression. These epigenomic changes may lead to adverse health effects for exposed individuals and their offspring. While the development of epigenomic biomarkers for exposures could facilitate understanding of these exposure-related health effects, such testing could also provide unwanted information.

**Objectives:** Explore Veterans’ attitudes toward epigenomic biomarker research and the potential to test for past exposures that could pose intergenerational risk.

**Methods:** Semi-structured interviews with Veterans (n=22) who experienced potentially harmful exposures during their military service.

**Results:** Twenty Veterans said they would hypothetically want to receive epigenomic information related to their toxic exposures and potential health impacts as part of a research study. Veterans identified nine potential benefits of this research, including promoting insights concerning intergenerational health, identification of early health interventions to mitigate the impact of exposures, and additional knowledge or explanation for their experiences. At the same time, 16 participants noted potential risks, including psychological distress in response to results, concerns about receiving non-actionable, uncertain, or inaccurate results, and issues related to privacy and discrimination. Ten participants also identified at least one condition in their children that they thought could be related to their exposure and most said they would be interested in receiving research results related to their children’s and grandchildren’s risk of developing a health condition associated with their exposure.

**Discussion:** Results suggest that Veterans might welcome benefits of epigenomic research related to military exposures yet have some concerns about potential negative impacts.

## Introduction

Several generations of U.S. military Veterans have faced exposure to a variety of toxic substances and environments during their time in service, and these exposures are often associated with poor health outcomes. For example, Vietnam era Veterans who were exposed to chemicals such as Agent Orange experience elevated rates of chronic respiratory conditions, heart disease, hypertension, and other ailments.^1^ Between 25-32% of Gulf War Veterans suffer from Gulf War Illness, a condition marked by a number of symptoms that are closely linked with chemical exposure.^2^ Veterans of the Global War on Terrorism who were exposed to hazards including burn pits and improvised explosive devices in Iraq and Afghanistan show an elevated risk for conditions such as chronic respiratory diseases.^3^ Under the Biden administration with the signing of the Sergeant First Class Heath Robinson Honoring Our Promise to Address Comprehensive Toxics (PACT) Act of 2022, there is great interest and investment in studying these exposures and their health impacts.

One potential way to address this is through the identification of epigenomic biomarkers associated with exposures. There is a growing body of literature suggesting that toxic exposures such as those encountered during military service may result in epigenomic changes and those changes may be linked to adverse health effects for the exposed individual and their offspring.^4^ Alterations in DNA methylation have been associated with a number of environmental factors, including chemical exposure and air pollution.^5^ One study of reproductive health among female Gulf War Veterans indicated that 38% percent of participants reported developmental disorders in their children, and the use of pesticide cream during deployment was associated with higher odds of these disorders being present.^6^

The development of epigenomic biomarkers of exposures could enable studies on the associated health effects that could lead to earlier and more accurate detection of disease, prediction of future health impacts, preventive and risk mitigation measures, and specific treatments through the identification of causal factors of disease.^7^ Clinical trials in oncology have shown that epigenomic biomarkers have both prognostic and predictive value, and epigenomic modifications can be used to assess the effectiveness of therapeutic interventions.^8^ Additionally, epigenomic biomarkers may allow for diagnosis and targeted therapy to occur simultaneously.^9^ The results of epigenomic testing, however, may also provide information that is distressing or burdensome to some individuals, particularly in the form of incidental, non-\actionable, or heritable results.^10^ These concerns may affect both the exposed individual and possibly their offspring. Increased concerns over privacy and discrimination when compared to traditional genetic testing may also be present, as it is uncertain whether genetic non-discrimination laws apply to epigenomic data.^11^

To develop epigenomic biomarkers for toxic exposures experienced in military settings, Veterans would need to be willing to participate in this research. While it is currently unknown what Veterans think about research on epigenomic biomarkers, several studies have shown that the Veteran population exhibits a high degree of willingness to participate in health research, especially when the research has the potential to help other Veterans.^12-14^ Veterans have also been found to generally have a positive attitude toward genetic research. One study of Veterans Affairs patients found that 83% of respondents believed that a database of genetic information and medical records for research purposes should be established, and 71% said they would definitely or probably participate.^14^ A separate study examining attitudes toward receiving genetic research results found that over 90% of Veterans would like to receive results, and there was no difference in attitudes between Veterans and non-Veterans.^15^ However, there is a paucity of research specifically regarding attitudes toward epigenomic research, and the data that does exist suggests that the general public has a limited understanding of the field.^16^

To better understand Veterans’ attitudes toward research on epigenomic biomarkers for exposures, we conducted qualitative interviews with Veterans who experienced potentially harmful toxic exposures during their military service. Here we report their interest in and anticipated benefits and risks of epigenomic research and testing, as well as their attitudes toward intergenerational risk testing.

## Methods

### Participant Recruitment

Veterans who sought medical attention for military exposure-related concerns through formal Department of Veterans Affairs’ registry examinations (Agent Orange, Gulf War, Airborne Hazards and Open Burn Pit (AHOBP) registries) were identified from medical records and eligibility and contact information was abstracted in accordance with a partial waiver of informed consent and HIPAA. Each registry has specified eligibility criteria primarily related to military deployment location and time period (Agent Orange-Vietnam 1962-1975; Gulf War-Persian Gulf region and Southwest Asia 1990-present; AHOBP Southwest Asia or Djibouti 1990-present or Afghanistan 2001-present). The registries are voluntary, and Veterans generally must seek out a registry examination, i.e., they were not often referred by a treating clinician.^17^ Veterans who completed a clinical encounter for one these registries at a single, large VA medical center between 2020 and 2021 were mailed an invitation letter to participate in the interview study and instructions on how to opt out of an invitation phone call. Ten business days after mailing the letter, a trained research coordinator phoned the Veteran at the phone number listed in the medical record and explained the study, confirmed eligibility, and invited the Veteran to participate. Three phone attempts were made. Interested Veterans were scheduled for a semi-structured interview.

### Interviews

Semi-structured interviews were conducted in late 2021 with Veterans via phone or video conference by a trained qualitative researcher (SP or CA). Interviews began with verbal informed consent as approved by the Institutional Review Board (IRB). The interviewer explored the Veteran’s experience with toxic exposures during their military service and the perceived impact on them and their families. The interviewer then elicited Veterans’ views toward hypothetical research and testing for epigenomic biomarkers for past toxic exposures. Interviewees were told that researchers would like to conduct studies to learn if a blood test could be developed to determine whether they had experienced a toxic exposure and identify what health risks they and their offspring might have due to that exposure. They were then asked what they thought the benefits and risks of such research would be and whether they would be interested in receiving results from such research for themselves and for their children and grandchildren. See supplementary materials for interview questions. Interviews were recorded and transcribed verbatim. Interviewees were invited until we reached thematic saturation^18^ on anticipated benefits and risks of epigenomic testing for toxic exposures. Demographic characteristics were self-reported and included to characterize the sample.

### Analysis

All transcripts were analyzed by two authors (SP and CA) aided by MAXQDA (VERBI Software, Berlin) to identify salient themes in the data.^19^ The initial broad deductive approach focused on participants’ perception of the impact of their exposure, their interest in hypothetical testing for epigenomic biomarkers for themselves and their families, and their perceived benefits and risks of such testing. The second step used an inductive approach to abstract salient themes within the broader codes.^20^ Any discrepancies in codes or abstractions were identified and discussed until consensus was reached. Illustrative quotes include participant number and cohort (i.e., Agent Orange, Airborne Hazards & Open Burn Pit, or Gulf War).

The research was approved by IRB and other oversight committees at Michael E. DeBakey VA Medical Center and Baylor College of Medicine (protocol H-49702).

## Results

### Participants

One hundred and twenty Veterans who sought medical attention for military exposure-related concerns were mailed an invitation letter to participate in an interview. Twenty-two Veterans were interviewed (Table 1) and 15 additional Veterans were reached by phone and either declined to participate or were unresponsive to scheduling attempts after the initial phone contact. Twenty of 22 Veterans said they would hypothetically want to receive epigenomic information related to their toxic exposures and potential health impacts. As one Veteran described,

**Table 1.** Participant Characteristics by military cohort/registry participation.

|  | **Agent Orange**<br>**(n=8)** | **Gulf War**<br>**(n=7)** | **Airborne**<br>**Hazards &**<br>**Open Burn Pit**<br>**(n=7)** | **Overall**<br>**(n=22)** |
| --- | --- | --- | --- | --- |
| **Age (years)** | 70.4 ± 7.3 | 55.6 ± 7.4 | 43 ± 7.8 | 55.7±13.5 |
| **Gender** |  |  |  |  |
| Male | 8 (100%) | 6 (86%) | 6 (86%) | 20 (91%) |
| Female | 0 | 1 (14%) | 1 (14%) | 2 (9%) |
| **Race/Ethnicity** |  |  |  |  |
| White | 5 (63%) | 3 (43%) | 6 (86%) | 14 (64%) |
| Black or African American | 3 (37%) | 3 (43%) | 1 (14%) | 7 (32%) |
| Multiple | 0 | 1 (14%) | 0 | 1 (4%) |
| **Education** |  |  |  |  |
| High School | 3 (37%) | 1 (14%) | 4 (57%) | 8 (36%) |
| Some College | 1 (13%) | 4 (57%) | 0 | 5 (23%) |
| Bachelor's Degree or Higher | 4 (50%) | 2 (29%) | 3 (43%) | 9 (41%) |
| **Biological Children** |  |  |  |  |
| Yes | 8 (100%) | 6 (86%) | 7 (100%) | 21 (96%) |
| No | 0 | 1 (14%) | 0 | 1 (4%) |
| **Grandchildren** |  |  |  |  |
| Yes | 3 (37%) | 1 (14%) | 2 (29%) | 6 (27%) |
| No | 5 (63%) | 6 (86%) | 5 (71%) | 16 (73%) |

> Why anybody would not want to know that, because it could give you such peace of mind. In my opinion, it could give you such peace of mind at knowing what’s going on with you, what possibly contributed to it, and whether or not it’s something that is going to be passed down to your kids. I would love to know that information and if I could get that from a single blood test. Holy cow, where do I sign up? (*7BP, Airborne Hazards & Open Burn Pit*)

Their reasons for wanting this information are captured in the anticipated benefits below. For the two Veterans who indicated they would not want to receive epigenomic information, one felt he could not handle any additional information and noted that he did not want to know about his risk for any future health problems: “Hopefully […] all my health problems are stopped. I don’t want to know of more new ones coming down the road,” (*8AO, Agent Orange*). The other was unsure and explained that his concern was that the information would not be actionable.

### Anticipated Benefits

Veterans anticipated several benefits of research on epigenomic biomarkers for military exposures. We identified nine distinct benefits, and most were related to the potential for return of individual research results. Most commonly, Veterans noted the potential of such information to **promote intergenerational health** (n=12). One Veteran explained why he would want to receive information about intergenerational health risks associated with his military exposures, “Yeah, because I’d like to tell my daughter […], hey, I’ve had this problem. I’ve been exposed to this. This might be an issue for you later,” (*6BP, Airborne Hazards & Open Burn Pit*). Another Veteran explained that knowing how his exposures could impact his children would be helpful:

> Well, because I would want to know how would it impact them, what kind of lifestyle are they going to have if they going to have one at all, are they going to be crippled, blind or some kind of physical defect, how is this going to affect them? So yeah, I would want to know in advance. Maybe if something can be done before that ever happens, maybe they can fix it. (*1AO, Agent Orange*)

The second most commonly anticipated benefit of this research was the potential to provide important information that could lead to **early health interventions to mitigate the impact of exposures**, “I think the main thing of knowing information like that would be doing screening you might not otherwise be doing. I mean, if I knew something might be cancer causing on down the line, then I certainly would want them to be able to catch that early,” (*1GW, Gulf War*). Another Veteran explained:

> Well, like with Agent Orange, sometimes it was 20 or 30 years before the effects of it, they got the cancer or something. So, if they had the blood test, then they could see maybe there are treatments or medications or stuff that might help, might delay… I don’t know, I’m not a doctor, but it seems like that would be real good. (*7AO, Agent Orange*)

Seven Veterans felt that information pertaining to epigenomic effects of military exposure would be beneficial simply for the sake of having **additional knowledge or an explanation** for their experiences, even if it didn’t lead to a specific intervention, “I think the information, if we look at it the right way, would be helpful and I would want to know. Even if it wasn’t anything that was life threatening, at least it would maybe help me to understand why I’m dealing with some of the things I’m dealing with,” (*2GW, Gulf War*).

Similar to the Veterans who felt that epigenomic risk information could lead to early health interventions, some (n=6) felt that such information could **inform treatment** for their current health concerns and symptoms:

> If you could fix what’s going on with me having to all of a sudden now in my forties and fifties carrying an inhaler because all of a sudden I’m asthmatic during allergy season… Yeah, so I don’t have lung infections and all those fun things. Yeah, I would think if it improved my health, because we could do something about it, I would be happy. (*3BP, Airborne Hazards & Open Burn Pit*)

Five Veterans explained that predictive health risk information could help them with **life planning**:

> How would I use [predictive health risk information]? I would use that information to plan for the eventuality of, all right, if I’m at risk for this, my life expectancy could be this. I will know that at this point, if I haven’t had these things done by then, I need to get them done at this point before the end comes along. It would help me to prepare for my life ending and my family, or at least my son being able to continue with his life and move forward and not have to be so much of a burden. It’s better to know what you may go from, in my opinion, than not knowing because then you can’t really prepare for it. (*7BP, Airborne Hazards & Open Burn Pit*)

Only a few Veterans (n=3) noted that epigenomic research results could prove that they had experienced a toxic exposure and thus **facilitate obtaining disability benefits** related to that exposure, “It would all be good for my benefits and compensation from the Gulf War area, yeah,” (*3GW, Gulf War*). And two of those three noted specifically that it could help their children or grandchildren obtain benefits if they were impacted by their exposure:

> Well, I mean, let’s say they got the perfect solution, and they said, “Yep. This is what we found. And it’s proven based off of all of these data points.” Then what it provides is, you can’t deny it. If it’s a positive test… Like a COVID test, if it’s a positive test, you got it. There’s no denying it. So, if you can find a test that actively says, “Yes, you were exposed to these hazards because of your deployment operations, and it will cause these issues for your health, your kids’ health, your family’s health, throughout the line, 100%,” then the VA has no way around it. They have to say, “Yep. You’ve got it. Here’s your disability. We’ll continue to provide medical support for the rest of your life,” or for however long they determine it’s necessary. (*5BP, Airborne Hazards & Open Burn Pit*)

Finally, there were three benefits noted by one Veteran each: that such a test could **provide peace of mind**, that knowing about intergenerational health risks caused by a parent’s exposure could **reduce self-blame** by letting children know their health conditions are not their fault, and that **participating in research** was a benefit in and of itself.

### Anticipated Risks

When asked about what risks might be associated with research on epigenomic biomarkers for military exposures, six Veterans noted they perceived **no risk**, “Oh, I just don’t see why anybody would object to it if it’s something that’s going to maybe let them know if they have a potential health problem or a future health problem. What would be the drawback to it?” (*7AO, Agent Orange*). Four of the six Veterans who felt there were no risks were from the Agent Orange cohort, while the other two were from the Burn Pit and Gulf War cohorts.

The remaining 16 Veterans named 13 different risks they thought could be associated with research on epigenomic biomarkers. Again, most anticipated risks were related to the receipt of individual research results. The most commonly cited risk was the potential for **psychological distress** (n=7). Veterans discussed how learning predictive health information associated with their exposure could cause stress, anxiety, fear, and depression, “Well, [learning health risk] could throw someone into a spiral,” (*5GW, Gulf War*). And yet, many Veterans also noted that although they recognized this as a risk of learning the information, they felt they themselves could handle it, and that the potential benefit of this information outweighed this risk:

> If you take me and you stand me up next to an average civilian 50-year-old, I think I’m in way better health, but if you take a guy like that and you go, oh, well, you’ve been exposed to this and you might have this problem and this problem and this problem, mentally that might affect people. There could be some negative things to that […] people might kind of take it as a prophecy […] “Hey, you were exposed to depleted uranium and it shows in your blood work and these are the symptoms for it.” Now that’s going to be in the back of my head forever. Oh, when am I going to get sick from this or… but I think knowing what you’ve been exposed to and knowing the symptoms of it or what could possibly happen is better than not knowing. (*6BP, Airborne Hazards & Open Burn Pit*)

The remaining 12 risks were noted by only one or two Veterans each. Several of these anticipated risks stemmed from perceptions of the types of results one could receive as part of the research study or concerns about the accuracy of those results. These risks included the potential to receive **non-actionable information** (n=2) or **uncertain results** (n=1), the risk of **misdiagnosis** (n=2), and the potential for **overreliance on testing** (n=1) to the exclusion of other relevant information.

Another group of risks described centered around privacy and potential for discrimination. Veterans named concerns about **confidentiality risks** (n=1), as well as the risk of **insurance discrimination** (n=2) or **employment discrimination** (n=1):

> I’m probably willing to do that now, for example. But when I got right out of the service and I’m trying to get a job and I got to have my flight physical and I got to have all that and this might show a marker where, “Oh, you’ve been exposed to this, which might lead to that,” and as a result, what is that going to do to my insurance? I’m not opposed to it, but I wouldn’t be a volunteer when I was flying. (*3AO, Agent Orange*).

Of note, this Veteran went on to talk about how he would have felt comfortable getting such testing while in the military given the protections in place, but thought the risk of employment discrimination was higher in the civilian context:

> If I was in the military at the time and that would have happened [epigenomic testing offered], I would more than likely would’ve taken the test. It’s just that when I got out of the military, now you’re in a whole different environment. The civilians are not like the military. Once you’re in there, the military understands the environment that you’re in and stuff like that, so I would hope at that point in time that adaptations would be made and they would understand, so I would not be against it. Now, go out into the civilian world, which is totally different than the military. Then, in some ways I think the civilian world is more cutthroat than the military. (*3AO, Agent Orange*)

Other Veterans noted lifestyle or family-related risks, including a risk of this epigenomic risk information leading to **poor lifestyle choices** if they thought that the health condition they were at risk for was inevitable (n=1), having a **negative impact on an exposed Veteran’s child** (n=1), or the potential for this information to **deter exposed Veterans from having children** (n=2) they might otherwise have wanted, “Well, you can’t have kids, man, because you’re going to cause them to have all these problems,” (*5GW, Gulf War*).

Finally, three Veterans described two risks specifically related to the military service context. First, two Veterans noted that this information could be used to **block a Veteran from accessing benefits** (n=2) to which they might otherwise be entitled, “No [cannot think of any risks], I mean, unless somebody’s maybe putting in a claim for something saying they were exposed to something, but then the blood test proves they weren’t, then they don’t get compensated,” (*2GW, Gulf War*). One other Veteran suggested that linking toxic exposures in military service to health problems could **deter military service** (n=1), “If anything will come up, and if so, come up out in that risk, then my kids ain’t going nowhere when I finish, ain’t let them go to the military at all. […] I can’t see any other [risk], besides that it’d be lack of military personnel,” (*6GW, Gulf War*).

### Perception of Intergenerational Impact and Attitudes toward Testing

We also discussed Veterans’ perspectives toward epigenomic research that could inform them about health risks to future generations stemming from their exposures. When asked whether they had any concerns about their exposures impacting their children or grandchildren, 10 participants identified at least one health condition in their children they thought could be associated with their exposures. Veterans named conditions including premature birth, learning disabilities, neurological symptoms including seizures and sleepwalking, heart and lung defects, bipolar disorder, and type 1 and type 2 diabetes. According to the interviewees none of these conditions had been formally linked with the Veteran’s exposure. When asked if they would be interested in learning their children’s and grandchildren’s risk of developing a health condition associated with their exposure, 19 participants reported they were interested, and one was unsure. Of the remaining two participants, one did not have children and the other had children who were born before the Veteran’s service and exposure. Veterans’ anticipated benefits and risks of testing for intergenerational risk are included in the sections above, as many themes of risk and benefit were discussed together and not necessarily distinguishable between attitudes toward learning risks for themselves versus their offspring.

## Discussion

Epigenomic biomarkers could identify past toxic exposures and predict future health impacts for exposed Veterans and their offspring. Yet, such testing could also provide unwanted information or have negative effects on those who have experienced exposures and their offspring. Our results suggest that Veterans who have experienced potentially toxic exposures during their military service are generally in favor of this hypothetical research and anticipate benefits, especially benefits associated with individual results. Even though most Veterans noted some concerns about the possibility of unintended negative impacts, nearly all Veterans indicated they would be interested in receiving epigenomic information about themselves and their children and grandchildren.

Like other studies of Veterans’ perspectives toward research participation, we found a wide range of reasons why Veterans may choose to participate. Other studies have also found that Veterans are motivated to participate in research by the potential to learn about the causes of their health issues,^13-15,21,22^ but this is not always the primary motivation.^13,14^ One study of Iraq- and Afghanistan-deployed US Veterans found that main motivations to participate in health-related research were adequate compensation, desire to help other Veterans, and the perceived significance of the research.^13^ It is possible that the potential to help other Veterans was less commonly cited as an anticipated benefit in our work because our interviewees focused on the potential to receive research results about their individual epigenomic risk profile. Given our interviewees’ attitudes toward helping other Veterans in other sections of the interview (e.g., when discussing desired reparations for toxic exposures, these data analyzed and reported separately), it is possible that we would find similar motivations to help other Veterans if we had described in more detail the potential for gaining scientific knowledge and group benefits.

Our findings about Veterans’ anticipated risks of participating in this hypothetical epigenomic research also echo findings in the literature. The potential for psychological harm in response to learning health risk information, the most common theme of risk we found, has long been a concern about genetic testing and genetic research across many contexts.^23-27^ The remaining themes of anticipated risks identified by our participants were not frequently cited, but paralleled the types of barriers to participating in health research identified by Veterans in other studies. These included concerns about privacy and confidentiality, and distrust with federal institutions.^13,22^ Our findings, overall, suggest that Veterans’ attitudes toward epigenomic research are similar to their attitudes toward other types of health and genetic research.

It is important to interpret our results within the context of our study. First, our study reports on the perspectives of Veterans who have experienced potentially toxic exposures. It is possible that Veterans who have not experienced such exposures and subsequent negative health effects would be less positive toward this research and perceive a different risk to benefit ratio. Whereas almost all of our participants reported they would want to receive epigenomic information related to their exposures because they felt the potential benefits outweighed those risks, those who have not experienced exposures may feel those risks outweigh the potential benefits. One study, however, did find that exposure to hazards during one’s service was not associated with willingness to hypothetically participate in a large database of genetic information and medical records for research purposes,^14^ though the hypothetical research described in that study did not focus specifically on exposures. Second, to discuss epigenomic research in lay terms, we described it as research to develop a blood test that could tell whether someone had been exposed to certain chemicals, whether someone might develop health problems from those exposures, and whether that risk could be passed on to children or grandchildren of people who were exposed. As such, it is possible that some of our participants were imagining a truly diagnostic test that could determine exposures and health risks with certainty instead of a test that could identify probabilistic risk. Given our interviewees’ focus on the potential to receive individual results, research in this area should be careful to avoid therapeutic misconception. Finally, like all qualitative research, our findings are not meant to be generalizable, but rather transferable to similar contexts.^28^ The participants in this study were all users of the Veterans Health Administration (VHA) and had participated in an established registry examination; approximately half of all living Veterans have ever used VHA services and a smaller fraction of eligible Veterans participate in these registries.

Given the recent investments via the PACT Act in addressing toxic exposures experienced during military service, there is great interest in research that could identify biomarkers associated with exposures and resulting health impacts. Our qualitative study demonstrates that Veterans who have experienced potentially toxic exposures during their military service are generally positive toward research on epigenomic biomarkers for exposures, especially if their concerns about unintended consequences can be addressed. These findings can be used to design research studies that anticipate the hopes and concerns of Veterans who may be asked to participate in such research, which can reduce barriers to enrollment, but also help set realistic expectations for participating.

## Data Availability

Aggregated qualitative data are available from the corresponding author upon reasonable request in accordance with VA privacy requirements.

## Acknowledgements

We thank Asha Richards and Saurendro Ghosh for their research assistance on this project.

## Funding

Funding for this research was provided by the Gulf Coast Center for Precision Environmental Health (NIH P30 ES030285) through a National Institute of Environmental Health Sciences Center Pilot Project grant. This research was also supported in part by the U.S. Department of Veterans Affairs, Veterans Health Administration, Health Services Research and Development Services at the Center for Innovations in Quality, Effectiveness and Safety (IQuESt; CIN 13–413), Michael E. DeBakey VA Medical Center, Houston, TX.

## Notes

Conflicts of Interest The authors declare they have nothing to disclose.

### Competing Interest Statement

The authors have declared no competing interest.

### Author Declarations

The IRB of Baylor College of Medicine gave ethical approval for this work.

